# Inferring pathogen superspreading potential using early spatial spread patterns

**DOI:** 10.1101/2025.10.28.25338965

**Authors:** Qing Yao, Renquan Zhang, Tom Britton, Sen Pei

## Abstract

Superspreading driven by individual variation in transmissibility shapes novel pathogen emergence and the effectiveness of control measures. Current approaches to estimating superspreading typically rely on cluster size distributions informed by contact tracing or genomic data, limited by data availability and potential sampling bias. Here, we examined the impact of superspreading on the early spatial spread of novel pathogens using a branching process model incorporating inter-county mobility in the United States (US). To represent individual transmission heterogeneity, we modeled the number of secondary infections using a negative binomial distribution with a mean reproduction number *R*_0_ and a dispersion parameter *r* that quantifies superspreading potential. Simulations suggest that stronger superspreading tends to slow early spatial invasion, with a larger variation in epidemic growth between different realizations. Using a graph neural network designed for epidemic inference, we demonstrated that *r* can be reliably inferred from early spatial spread patterns, robust to the spatiotemporal variation of *R*_0_ and case underreporting. Application to early COVID-19 data in the US revealed strong superspreading prior to nationwide lockdown (*r* = 0.50, 95% CI: [0.23 - 1.20]), followed by weaker superspreading afterward (*r* = 1.3, [0.64 - 3.18]). Our study offers a new approach to quantifying pathogen superspreading potential using population-level observations.

## Introduction

Superspreading, where a small proportion of individuals infect the majority of secondary infections, is a common feature of disease spread caused by individual variation in transmissibility^1–5^. Superspreading events have been reported for a range of pathogens including SARS^6–9^, measles^10,11^, Middle East respiratory syndrome coronavirus (MERS)^3,12^, and more recently, SARS-CoV-2^5,13,14^. Previous studies indicate that superspreading has a profound impact on disease transmission dynamics, particularly during the early stage of outbreaks^15–17^. Given the same reproduction number, diseases with strong superspreading are less likely to maintain sustainable transmission chains; however, once established in a population, outbreaks tend to be more explosive^1^. Such epidemiological characteristics can greatly impact the effectiveness of intervention measures^18–20^ and herd immunity thresholds^21–24^. As a result, estimating the superspreading potential of pathogens is crucial for epidemic control.

The branching process model is a convenient mathematical framework for modeling superspreading^25^. In this model, the number of secondary infections, or offspring, generated by one infectious person is modeled by a negative binomial distribution *NB*(*R*_0_, *r*), where *R*_0_ is the basic reproduction number (the mean of the distribution) and *r* is the dispersion parameter. The parameter *r* quantifies the overdispersion of the offspring distribution: the variance of the distribution is 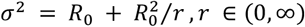. A smaller dispersion parameter *r* represents a more heterogeneous distribution of secondary infections, thus a stronger superspreading potential^1,26^.

Much effort has been made to infer the dispersion parameter *r* using outbreak data. One of the mainstream approaches relies on the distribution of transmission cluster size^27–29^, estimated using either individual-level contact tracing records^1,30,31^ or the classification of local versus imported cases^32^. More recent studies have utilized genomic data, reconstructing transmission trees to infer offspring variation^33,34^. However, high-quality contact tracing and dense sequencing are not always available during the early phase of an epidemic, and both may suffer observational bias towards large-scale transmission clusters and miss smaller ones. In addition, the geographical disparity in genomic data collection may limit the representativeness of those datasets^35^. Other studies attempt to use incidence-based likelihoods, where observed cases are modeled as negative binomial random numbers^36,37^.

While more broadly applicable, this approach still faces several limitations, such as overlooking the spatiotemporal correlation of incidence in the likelihood and numerical instability, especially in settings with low case counts and short time window^36,37^. These challenges motivate the development of alternative approaches that leverage spatiotemporal patterns in more accessible population-level data.

The early spatial spread of a novel pathogen could be shaped by superspreading as transmission heterogeneity can impact the probability of stochastic extinction (or self-sustained transmission) when the pathogen is introduced into a new region. If validated, spatial spread patterns can offer an alternative dynamical signature to estimate dispersion parameters using population-level statistics. Metapopulation models informed by human mobility are widely used to simulate the spatial spread of infectious diseases^14,24,38–41^. However, integrating individual-level variation into these population-level models is not straightforward. In addition, metapopulation models are typically represented by a set of ordinary differential equations with exponentially distributed infectious period, which deviates from the actual distributions estimated using real-world disease data.

In this study, we developed a branching process model with a spatial structure connecting over 3000 counties in the United States (US). Through extensive model simulations, we showed that superspreading can introduce substantial uncertainty in the early spatial spread of novel pathogens and impact the invasion speed across locations. To solve the inverse problem of inferring superspreading potential from early spatial spread patterns, we designed a Bayesian last-layer graph neural network (BLL-GNN)^42–44^ that couples a deterministic graph-convolutional encoder with a Bayesian output layer. This approach is physics-informed^45–47^: inter-county mobility is encoded as the graph structure in the GNN, branching-process dynamics provide the prior knowledge that constrains the learning process, and the Bayesian layer quantifies parameter uncertainty. The inference framework was validated using extensive simulations of outbreaks under various transmission conditions. Results indicate that this method can accurately estimate pathogen superspreading potential using population-level observations, robust to the spatiotemporal variation of the basic reproduction number *R*_0_ and case underreporting. We applied the inference framework to early COVID-19 data in the US and estimated that SARS-CoV-2 possessed strong superspreading during the early outbreak, in agreement with findings from other studies^5^.

Additionally, the estimated superspreading dropped substantially after the nationwide lockdown, suggesting the potential effect of interventions on reducing transmission heterogeneity.

## Results

### Spatial spread patterns with superspreading

We developed a branching process model with a spatial structure to simulate the invasion of a novel pathogen across US counties. This model jointly incorporated a susceptible-exposed-infectious-recovered (SEIR) dynamics, individual variation in transmissibility, and human mobility between US counties (Methods). Specifically, the number of secondary infections produced by one infectious person was drawn from a negative binomial distribution *NB*(*R*_0_, *r*). Note, in statistics literature, the negative binomial distribution is usually parameterized using *p*, the probability of success in each Bernoulli trial, and the dispersion parameter *r*. In the context of epidemiology, the basic reproduction number *R*_0_ is linked to the parameter *p* through *R*_0_ = *r*(1 − *p*)/*p* (Methods). For interpretation convenience, we used *R*_0_ and *r* to parameterize negative binomial distributions throughout this study. An illustration of the model structure is shown in Fig. 1. By varying the basic reproduction number *R*_0_ and the dispersion parameter *r*, we can explore the impact of intrinsic disease transmissibility and superspreading potential on the early spatial spread patterns.

**Fig. 1.**
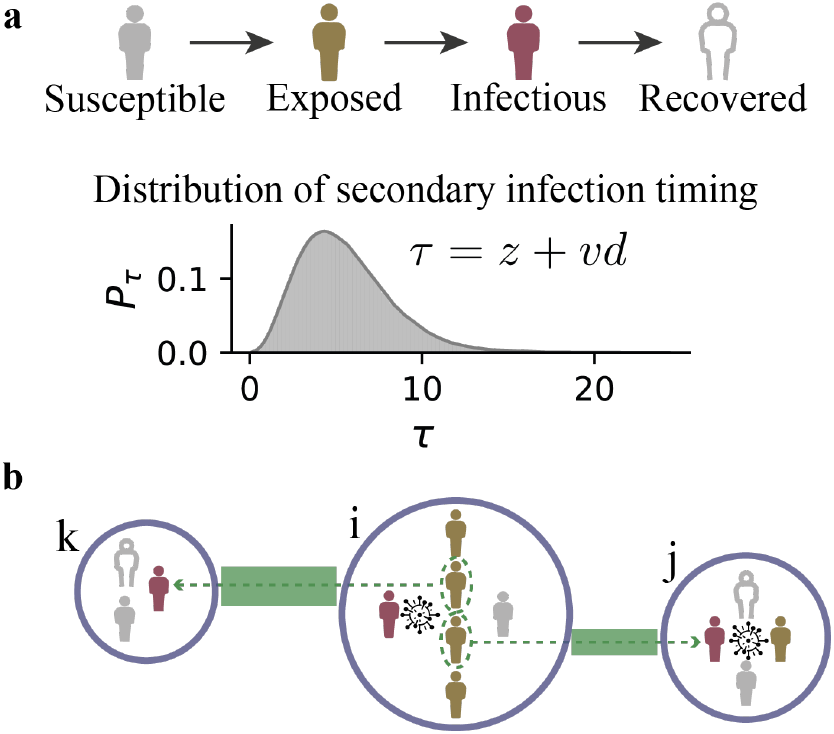
Illustration of the branching process model dynamics. (**a**). Transition of individual states in the model. A person becomes infected when transitioning from susceptible to exposed state. Each individual has a random latent period of *z* days following a Gamma distribution, after which it becomes infectious and recovers after an additional *d* days drawn from a Gamma distribution. An infectious individual infects *k* individuals, each uniformly distributed over the infectious period. The number of secondary infections *k* was generated by multiplying a negative binomial random number by the fraction of susceptible population in that location. The timing for generating a secondary infection is *τ* = *z* + *vd*, where *v* is drawn from a uniform distribution *U*[0,1]. The inset shows the probability distribution of *P*_I_. (**b**). A network diagram illustrating the spread of infectious disease across counties. In this diagram, each circle represents a county, and edges represent commuting flows between them. The grey, yellow, red, and hollow human icons represent susceptible, exposed, infectious, and recovered individuals, respectively. In county *i*, an infectious person (red) leads to four exposed individuals (yellow). Of these, two commute to counties *J* and *k*, respectively. In county *J*, the exposed individual becomes infectious (red) and further infects another person (yellow). In contrast, in county *k*, the exposed commuter becomes infectious (red) but does not result in any new infections.

We first examined the geographical dispersion of a novel pathogen after a certain period of transmission. We simulated outbreaks originating from New York County (Manhattan, NY) with various *R*_0_ and *r* configurations for 60 days, starting from 100 infected individuals (Methods). In each experiment, we assigned the same *R*_0_ and *r* in all counties. To capture the stochastic dynamics, 300 independent simulations were performed. We defined a county as “infected” if local daily new infections exceed 10 per 100,000 people. Taking average over all simulations, the number of infected counties by day 60 increased with growing *R*_0_ (Fig. 2a). For a fixed *R*_0_, stronger superspreading (i.e., smaller *r*) led to a lower number of infected counties on day 60, averaged across 300 independent simulations. Additional to lower average number of infected counties on day 60 for superspreading, another difference is that superspreading also increases variation (e.g. standard deviation) between simulations. (Fig. 2b).

**Fig. 2.**
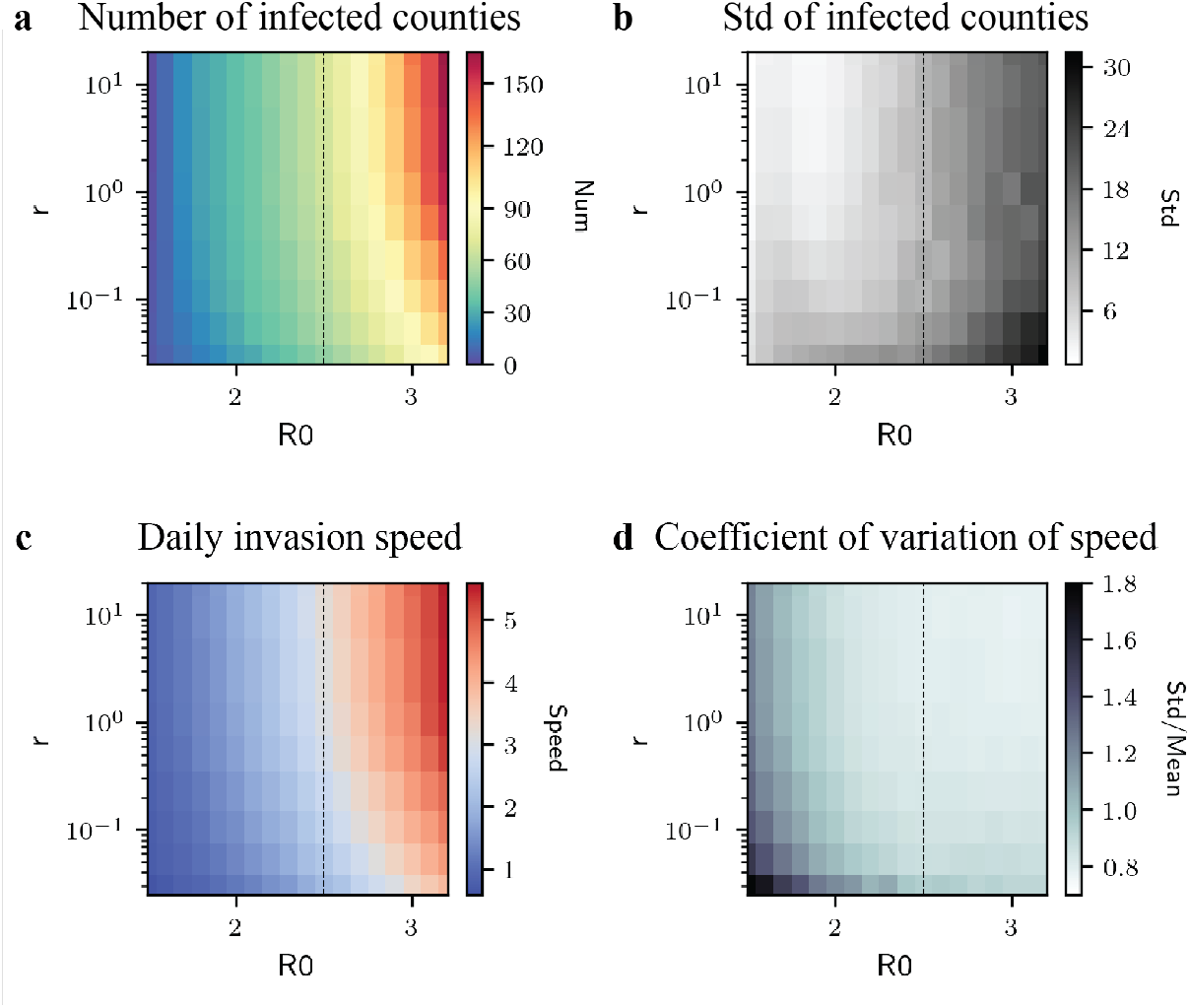
Spatial spread patterns of the branching process model. (**a**). The number of infected counties (with daily new infections at least 10 per 100,000 people) by day 60 of simulations using various combinations of *R*_0_ (1.5 ≤ *R*_0_ ≤ 3.3) and *r* (0.025 ≤ *r* ≤ 20). For each parameter combination, we performed 300 independent simulations and presented the mean number of infected counties. The vertical dash line marks the parameter *R*_0_ used in simulations shown in Fig. 3. (**b**). The standard deviation of the number of infected counties by day 60 for 300 independent simulations. (**c**). The daily invasion speed (i.e., the daily number of new counties with at least one infection) averaged over the 60-day period for outbreaks generated using different combinations of *R*_0_ and *r*. (**d**). The coefficient of variation of daily invasion speed within the 60-day simulation window, defined as the standard deviation divided by the mean of speed. The coefficient of variation quantifies the relative variation of invasion speed around the mean speed during the 60-day period. Results were computed using 300 independent simulations.

We quantified the spatial invasion speed of pathogens using the daily number of counties that have first local infections. For instance, an invasion speed of 5 counties per day means that the outbreak produced at least one infection in 5 new counties each day. Averaged over the 60-day period, the mean invasion speed increased with growing transmissibility (*R*_0_) and decreased for higher superspreading (Fig. 2c). To measure the temporal variation of the invasion speed, we computed the coefficient of variation (COV) of daily invasion speed within the 60-day simulation window, defined as the standard deviation of invasion speed divided by its mean. The COV quantifies the relative variation of invasion speed around the mean speed during the 60-day period. Pathogens with a lower *R*_0_ had a larger COV of invasion speed, suggesting a more pronounced acceleration of spatial invasion over time (Fig. 2d). Given the same *R*_0_, stronger superspreading led to a larger COV, possibly due to the higher uncertainty in simulation outcomes.

To further compare the temporal evolution of spatial spread at different superspreading levels, we compared the number of infected counties over the 60-day period for *R*_0_ = 2.5, using the same parameter settings and initial conditions but different dispersion rates (Fig. 3a). To capture the variation of spatial spread generated using the stochastic branching process model, we performed 300 independent realizations for each parameter setting. In general, the models with smaller *r* (stronger superspreading) produced slower spatial dispersal during the early phase of the outbreak. For *r* = 20, the realization with a median number of infected counties by day 60 infected 67 counties. In contrast, for *r* = 0.025, the median trajectory only infected 48 counties, 28.3% lower than the median outcome for *r* = 20.

**Fig. 3.**
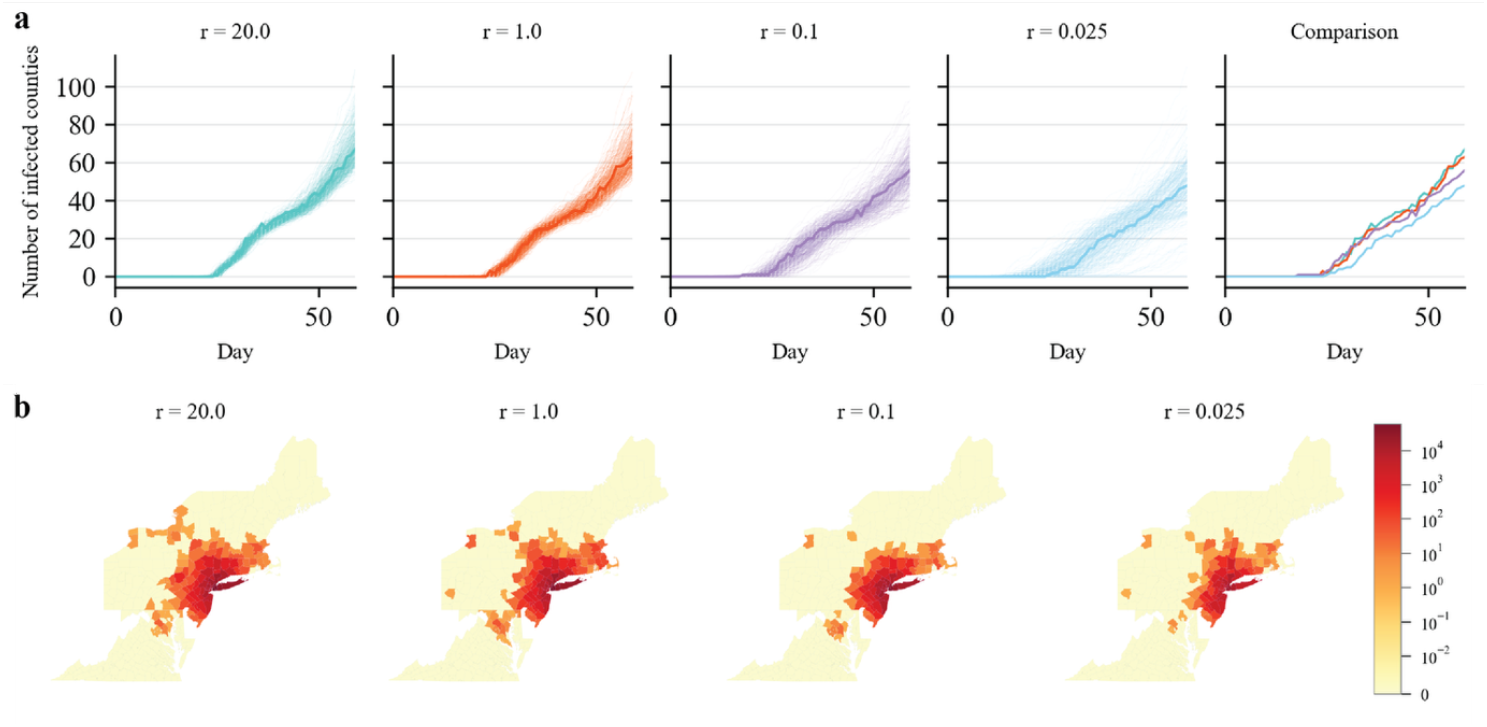
Effect of superspreading on spatial spread. **(a)** We compare the number of infected counties (daily new infections exceeding 10 per 100,000 people) over time for branching process models with *R*_0_ = 2.5 and varying dispersion parameters. A smaller *r* indicates stronger superspreading. For each *r* value, we performed 300 independent realizations. Thick solid lines show the trajectory for the realization that infected the median number of counties on T = 60 days. All the other realizations are plotted using thinner trajectories. The last panel compares the median trajectories for different dispersion parameters **(b)** Daily new infections in each county near the outbreak origin on day 60 of the simulation. Colors of counties indicate the daily new infections on day 60, generated using branching process models with different *r*. We visualize the outcome for the model realization that infected the median number of counties on T = 60 days (the thick solid lines in (a)). The outbreaks were initiated by 100 infections in New York County.

For branching process models, higher superspreading potential (*r* = 0.025) led to highly stochastic outcomes, indicated by the wide spread of trajectories generated from independent realizations, especially during the onset period between 15 and 25 days (Fig. 3a). For an outbreak with stronger superspreading potential, the initial spatial spread speed was more variable across different model realizations – some realizations could be faster due to occasionally more explosive infections, and some could be slower as transmission chains may die out. For *r* > 1, the effect of superspreading became less pronounced. We visualize the geographical range of infected counties in northeastern US on day 60 generated by branching process models with different *r* values (Fig. 3b). The branching model with large

*r*, whose offspring distribution is closer to a Poisson distribution, produced a faster spatial invasion for the pathogen originating from New York County during the 60-day period. Additional simulations with a higher *R*_0_ = 5.5 further revealed pronounced difference in spatial spread at the national scale among different dispersion parameters. (Methods, Fig. S1).

### The effects of stochastic extinction and superspreading events

Our simulations indicate that the effect of transmission heterogeneity on spatial spread has two folds. First, on average, a pathogen with strong transmission heterogeneity tends to spread more slowly into new areas during early outbreaks. Such slower spatial spread is primarily caused by the high extinction probability of an infection. For instance, for *R*_0_ = 2.5 and a dispersion parameter *r* = 0.025, the probability of stochastic extinction for one infection is 0.96 (Supplementary Information. Note, stochastic extinction probability is different from the probability of generating zero offspring, which is 0.89 here). In contrast, for *r* = 20, the extinction probability is only 0.12. For small dispersion parameters, the establishment of sustained transmission in a new location thus likely requires more introductions, which will take a longer time. Second, stronger superspreading potential leads to a larger variation in epidemic growth among independent model realizations. Some rare superspreading events may expedite spatial spread by producing a large number of seeds introduced into new regions, potentially offsetting the effect of high extinction probability of these introductions.

To better disentangle the effects of stochastic extinction and superspreading events, we performed simulations where the number of secondary infections was generated using a Poisson mixture distribution. Specifically, we replaced the negative binomial distribution in the branching process model with a Poisson mixture: *p*_*λ*_*Pois*(*R*_0_/2) + (1 − *p*_*λ*_)*Pois*(*λ*) for *λ* > *R*_0_, where *Pois*(*λ*) is a Poisson distribution with mean *λ*, and *p*_*λ*_ controls the mixture between the two Poisson distributions. To ensure the distribution mean to be *R*_0_, *p*_*λ*_ needs to satisfy *p*_*λ*_ = (*λ* − *R*_0_)/(*λ* − *R*_0_/2). By varying the parameter *λ*, we can adjust the extinction probability and the probability of superspreading events: a larger *λ* leads to a higher extinction probability and rarer but more explosive superspreading events. The effect of *λ* on the offspring distribution is thus similar to that of 1/*r* in the negative binomial distribution; however, its impact on extinction probability is less pronounced^48^. For *R*_0_ = 2.5, a large value of *λ* = 40 (*p*_*λ*_ = 0.97) leads to an extinction probability of 0.55 (Supplementary Information), much lower than the extinction probability for small *r* values in the negative binomial distribution. A comparison of extinction probabilities for negative binomial distributions and the Poisson mixture distributions is provided in Table S2.

We simulated outbreaks using Poisson mixture distributions with *R*_0_ = 2.5 and various *λ* values (Fig. S2). With a reduced difference in extinction probability, the median trajectories exhibited similar spatial spread speed irrespective of amount of superspreading events. However, we still observed an increased variance in the number of infected counties for larger *λ* values (Fig. S2), suggesting that superspreading events mainly contribute to the stochasticity of early spatial invasion dynamics.

### Inferring superspreading using Bayesian Last-Layer Graph Neural Network

As superspreading can affect the invasion dynamics of novel pathogens, the early spatial spread patterns may serve as a dynamical signature that allows the inference of superspreading potentials. However, this inference task is challenging for traditional approaches – for likelihood-based methods, it is difficult to handle the spatiotemporal correlation in case counts; for machine learning algorithms, it is hard to manually design features that capture superspreading potential. To address these challenges, we employed a graph neural network (GNN) framework, which can flexibly learn the nuanced spatiotemporal patterns associated with different levels of superspreading. GNN has been widely applied in spatiotemporal modeling of complex dynamical systems, including infectious disease transmission^49–51^. To further capture uncertainty in the inferred parameters, we extended the architecture with a Bayesian last-layer formulation. The resulting Bayesian Last-Layer Graph Neural Network (BLL-GNN) combines a deterministic graph convolutional encoder with a Bayesian output layer to estimate the dispersion parameter (Methods, Supplementary Information). This hybrid design enables efficient uncertainty quantification while maintaining scalability to large, time-varying mobility networks^42^.

We validated the inference framework using simulated outbreaks generate with different dispersion parameters. For a given reproduction number *R*_0_, we generated an outbreak using a ground-truth dispersion parameter *r*. Our goal is to estimate this ground-truth *r* using the simulated daily new infections obtained from one single realization. To train the BLL-GNN, we generated synthetic outbreaks using the same *R*_0_ and different *r* values as a training dataset. For each (*R*_0_, *r*) combination, we generated 180 independent outbreaks for training. The BLL-GNN was optimized using this simulated training data for the regression task to output dispersion parameters with the input of daily new infections in all counties. We then applied the optimized BLL-GNN to the simulated outbreak data to infer the ground-truth *r*.

To validate the performance of the neural network for diseases with different levels of transmissibility *R*_0_, we performed experiments for three fixed *R*_0_ values: 1.5, 2.5, and 5.5. For reference, a table for the basic reproduction numbers *R*_0_ and dispersion parameters *r* for a list of common infectious diseases are provided in Table S1. For each dispersion parameter, we generated 60 realizations of outbreaks and performed inference for each realization independently. In general, the graph neural network accurately inferred the dispersion parameter across a broad range of *r* values (Fig. 4a–c), except when *r* exceeded 10. This limitation is expected, as the negative binomial distribution converges to a Poisson distribution for large *r*, producing nearly indistinguishable spatiotemporal transmission patterns. Consistent with previous findings, estimation accuracy decreased once *r* surpassed unity, reflecting reduced overdispersion in the offspring^36,37^. The overall coverages of true *r* within the 95% CIs were 89.6%, 93.8% and 98.9% for *R*_0_ = 1.5, 2.5, and 5.5, respectively.

**Fig. 4.**
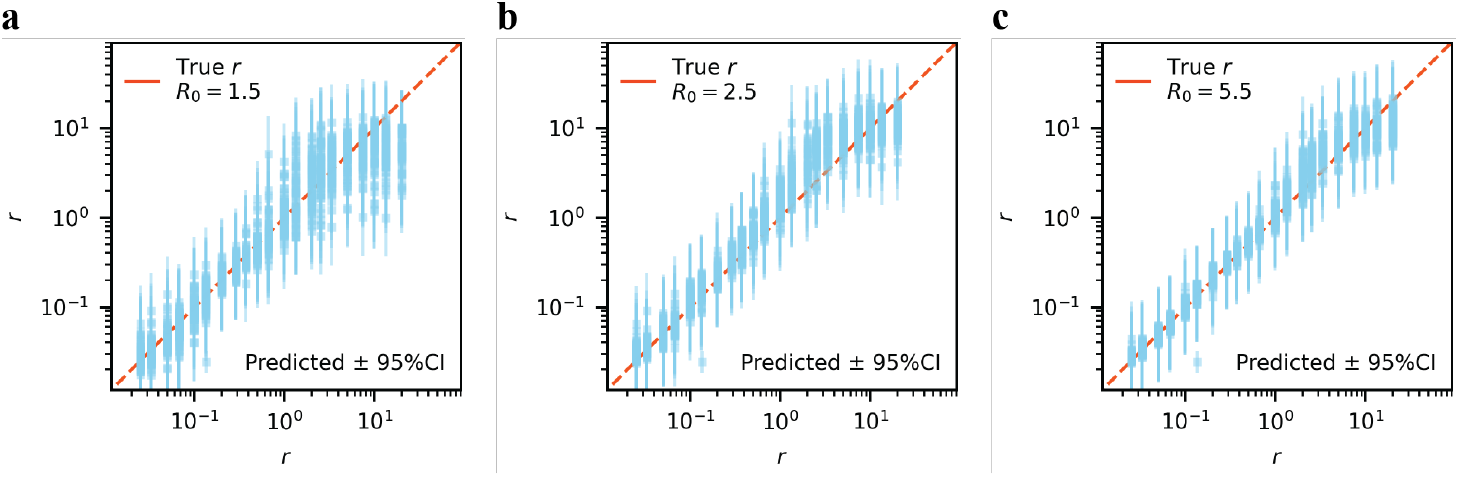
Performance of the Bayesian graph neural networks for synthetic outbreaks with different R0. We generated three groups of synthetic outbreaks using *R*_0_ = 1.5 **(a)**, 2.5 **(b)**, and 5.5 **(c)**. The x-axis shows the true values of the dispersion parameter *r* and the y-axis shows the inferred values for synthetic epidemics generated with different basic reproduction numbers *R*_0_. A perfect inference would lie on the dashed red diagonal line. For each true *r* value, we generated 60 out-of-sample independent realizations of the outbreak and performed inference for each realization separately. For each parameter *r*, the plot overlays inference results from 60 independent inference. Blue dots indicate the median point estimates and vertical bars represent the 95% CIs from the Bayesian model.

In real-world settings, only a single realization of the stochastic epidemic process is typically observed, with unknown reproduction numbers varying over time and across locations. To validate our approach under such conditions, we generated synthetic outbreaks with county-specific, time-varying reproduction numbers and tested the model’s ability to recover the true dispersion parameter (Fig. S3). For each county, we selected an initial *R*_0_ linked to population size and then gradually decreased *R*_0_ over time (Methods). To capture the general pathogen transmissibility, instead of estimating the reproduction number in each county, we estimated the crude daily effective reproduction number *R*_*t*_ at the national level using EpiEstim^52^, a statistical method based on renewal processes to estimate time-varying reproduction numbers. As the population infected during the early outbreak is negligible compared with the total population, we used the estimated effective reproduction number *R*_*t*_ as the proxy for the basic reproduction number *R*_0_. The estimated reproduction number matched the temporal trend of *R*_0_ in different counties (Fig. S4). We then used the estimated daily national reproduction numbers and different dispersion parameters to generate synthetic outbreaks as training datasets for the BLL-GNN (Methods), where the inputs are the daily case counts in all counties and the output is the dispersion parameter with associated uncertainty. To validate the trained BLL-GNN, we generated synthetic outbreaks using seven representative, ground-truth *r* values, among which three values were not used in model training and served as out-of-sample validation targets. Across the seven *r* values, the BLL-GNN accurately recovered the true dispersion parameters (Fig. 5a), despite that the reproduction numbers used in generating training datasets were different from the actual county-specific *R*_0_.

**Fig. 5.**
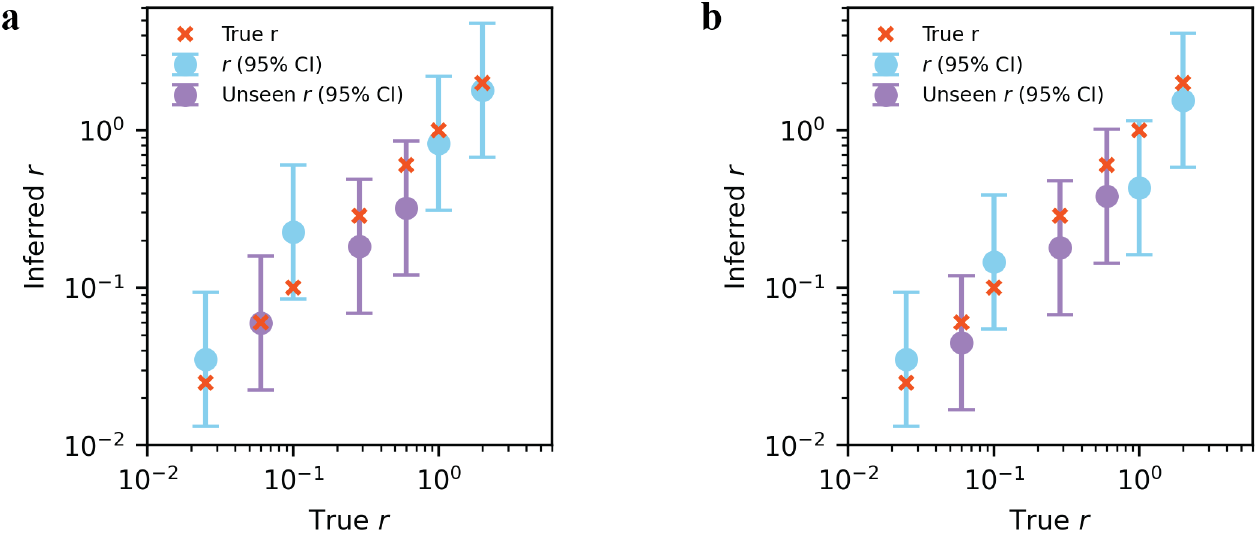
Performance of the Bayesian graph neural networks for outbreaks with location-specific time-varying *R*_0_ and underreporting. We generated synthetic outbreaks using reproduction numbers that vary over space and time and used the daily new infections (without underreporting) to infer dispersion parameters **(a)**. The x-axis shows the ground-truth *r* values used in epidemic simulation and the y-axis shows the inference results. The red crosses show the true *r* values. The dots indicate the median point estimates, and the bars represent the 95% CIs. Blue color shows the *r* values included in the generation of training datasets, and purple color shows the values not used in model training (i.e., out-of-sample validation targets). We additional included underreporting rates that vary in different locations and time and used underreported case counts to perform inference. Results are shown in panel **(b)**.

In the above experiments, we assumed all infections were captured by the surveillance system. In reality, underreporting poses a major challenge for real-world inference^24,53^. To examine the robustness of the inference to underreporting, we further incorporated underreporting in the simulated outbreaks. Specifically, we assigned an initial reporting rate for each county in the range of 10%-15% and then gradually increased it over time to 15%-20% (Methods). Using the underreported case counts, we applied EpiEstim to estimate the daily national reproduction number and found that the results captured the temporal trend of *R*_0_ (Fig. S4). For validation, we simulated outbreaks using seven ground-truth *r* values. Then we generated training datasets using the estimated reproduction numbers and various dispersion parameters, trained the BLL-GNN, and performed inference with the underreported case counts to estimate *r* (Methods). Even with severe, time-varying underreporting, the trained BLL-GNN was able to recover the true dispersion parameters (Fig. 5b).

### Estimating superspreading of SARS-CoV-2 in the US

We applied the inference framework to the early phase of the COVID-19 pandemic in the US, focusing on two periods: February 23rd–March 14th, 2020 (before the nationwide lockdown) and March 15th–April 18th, 2020 (after lockdown). Studies have shown that changes in population mobility are strongly associated with changes in disease transmission^54^. To better capture changes in population mobility during this period, we incorporated time-varying inter-county mobility constructed from anonymized foot-traffic data provided by Advan Research Corporation^55^ on Dewey Data platform^56^. In the inference, we used the daily new infections (both reported and unreported cases) estimated using a dynamic data-driven transmission model that accounted for time-varying ascertainment rates and reporting delays^24^. The estimated infection numbers were validated independently by serological data collected at different locations and time points^24^.

During the pre-lockdown phase, we estimated a dispersion parameter of *r* = 0.50 (95% CI: 0.23–1.20) (Fig. 6), consistent with a pooled estimate of SARS-CoV-2 superspreading in literature^5^. Such a low *r* value indicates a strong superspreading potential, as observed in several early COVID-19 clusters worldwide^5,13,14^. After the implementation of mobility restrictions and behavioral interventions, the inferred dispersion parameter increased to *r* = 1.3 (95% CI: 0.64–3.18), suggesting a noticeable reduction in transmission heterogeneity. This shift implies that control measures not only decreased the overall pathogen transmissibility but also reduced opportunities for large-scale superspreading events, possibly achieved by social distancing and limiting large gatherings.

**Fig. 6.**
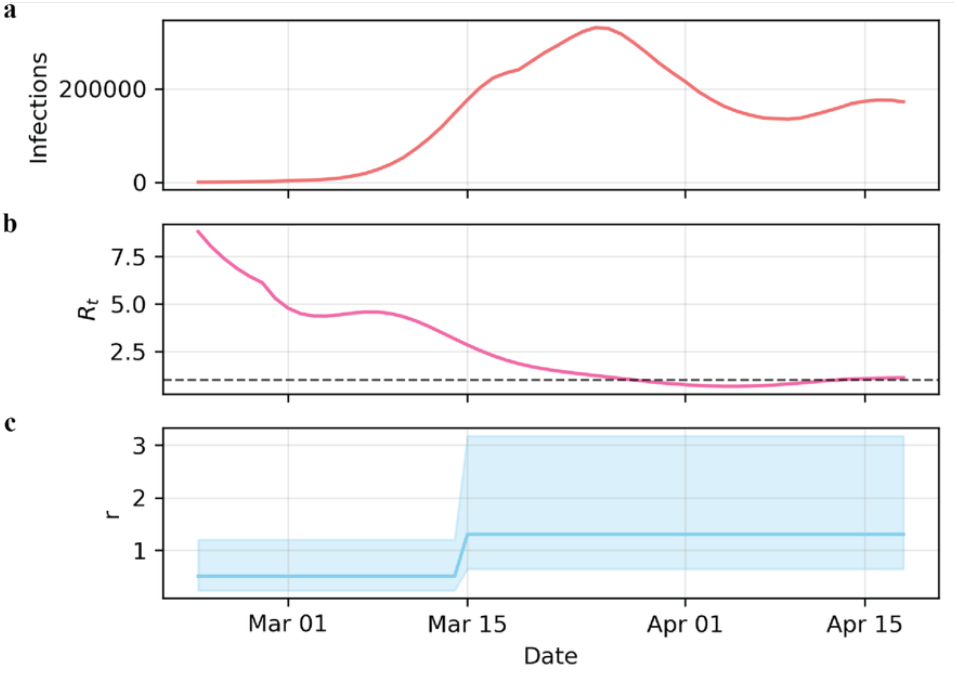
Estimating superspreading during the early COVID-19 outbreak in the US. Panel **(a)** Estimated daily infections (both reported and unreported) across U.S. counties from 23 February to 18 April 2020, from Pei et al^24^. **(b)** The time-varying national effective reproduction number reconstructed using EpiEstim (dashed line marks *R*_0_ = 1). **(c)** Estimates of the dispersion parameter *r* (smaller *r* indicates stronger superspreading) for the two periods: February 23rd–March 14th, 2020 (before the nationwide lockdown) and March 15th–April 18th, 2020 (after lockdown). Solid lines show posterior medians; shading areas show the estimated 95% CIs.

## Discussion

Transmission heterogeneity can impact the probability of establishing sustained transmission when a novel pathogen is introduced into a new region. Understanding how superspreading shapes the invasion dynamics of emerging outbreaks across locations is critical for epidemic control. We developed a branching process model with a spatial structure to explore how superspreading potential affects the spatial spread patterns of infectious diseases. Our simulations and the comparison with a Poisson mixture distribution reveal the separate effects of high extinction probability and rare superspreading events on spatial spread: a pathogen with a high extinction probability tends to spread more slowly into new areas during early outbreaks, while rare superspreading events lead to a large variation in the epidemic growth between different simulations. Predicting the spatial invasion speed for pathogens with strong superspreading potential is therefore challenging for a single model realization given this high stochasticity.

Our finding of the impact of transmission heterogeneity motivated us to infer the dispersion parameter using spatial spread patterns. Using population-level observations to infer individual-level epidemiological characteristics is advantageous in resource-limited settings where contact tracing and genomic data are not available. This offers an alternative approach to estimating dispersion parameters that complements existing tools. We demonstrated that the inference framework is robust to the spatiotemporal variation of pathogen transmissibility and underreporting, suggesting its potential practical utility in real-world settings. The inference results may support early assessment of controllability of emerging pathogens. Understanding this key epidemiological property is critical in informing effective interventions during the early stage of outbreaks.

Our study builds on recent advances in deep learning for stochastic physical systems and applies these ideas to epidemiology. Conceptually aligned with physics-informed and simulation-based machine learning frameworks, our approach leverages mechanistic knowledge from stochastic branching processes to guide neural-network inference. By representing inter-county mobility as a dynamic graph and integrating the statistical structure of branching processes into the learning framework, we demonstrate how deep learning can infer latent mechanistic parameters, such as the dispersion parameter, directly from observed epidemic data. This work illustrates how these principles and simulation-based inference can be applied to stochastic, high-dimensional systems such as epidemic spread.

Several limitations exist in this study. First, the branching process model does not fully represent the heterogeneity in real-world epidemic process. Multiple factors can contribute to superspreading, such as individual variation in susceptibility, infectiousness, and number of close contacts. In our model, all individuals are assumed equally susceptible, and the number of secondary infections for each infectious individual follows the same negative binomial distribution. This formulation represents heterogeneity in infectiousness rather than in social activity or contact structure, which can affect the risk of both infecting others and getting infected. Consequently, the model only captures random variation in how many secondary infections each case causes – for example, due to differences in viral load or chance attendance at large gatherings – but does not account for individuals who are consistently more socially active or at higher risk of exposure. Therefore, while we refer to strong overdispersion as superspreading, it should be interpreted as reflecting variability in infectiousness rather than the super spreader behavior. Future studies can further incorporate other factors contributing to superspreading. Second, to generate training datasets, we estimated the national reproduction number and ignored the variation of *R*_0_ across locations. The reproduction number was inferred using EpiEstim, which may yield overconfident estimates in the presence of overdispersion^57^ and has higher bias when cases are few^58^. A promising direction for future work is the joint inference of both transmissibility and superspreading within a unified framework. Lastly, the computational cost for running the branching process model increases substantially as the number of infections grows. Future studies should explore more efficient simulation techniques to speed up the algorithm.

## Methods

### Data

The U.S. Census cross-county commuting data were obtained from the US Census Bureau^59^. We used the 5-year commuting data to extract the population flow across the 3142 counties in the US. Population in each county was downloaded from the US Census Bureau Population and Housing Unit Estimates Datasets^60^. The mobile phone-derived foot-traffic data was provided by Advan Research Corporation^55^ through Dewey Data platform^56^. The estimated daily numbers of COVID-19 infections (both reported and unreported) in US counties were obtained from Pei et al.^24^

### Mobility networks

The static cross-county mobility network is constructed using the US Census cross-county commuting data. This dataset provides commuting patterns between counties, denoted as *n*_*i*j_, representing the daily number of people commuting from county *J* to county *i*. To remove negligible human mobility, we retained only commuting flows of more than 100 people per day. Subsequently, we normalized these flows based on the population of the originating county, ensuring they represent a proportion of the county’s total population. The resulting mobility matrix is denoted by *M* = {*m*_*i*j_}. The size of *M* is the number of all counties and county-equivalents in the US. Each entry *m*_*i*j_ for *i* ≠ *J* represents the probability of an individual commuting from county *J* to county *i*; *m*_*ii*_ represents the probability of an individual staying in the same county *i*.

The time-varying cross-county mobility networks in 2020 were built using anonymized foot-traffic data derived from mobile devices. The original dataset recorded the number of visits that anonymized users made from their home census block groups to various points of interest (POI) such as workplaces, restaurants and bars, and other public venues. To obtain county-level movement patterns, we aggregated these visits according to the county of residence and the county of destination POI. This produced a sequence of daily mobility matrices representing the number of trips between every pair of origin and destination counties from February 23 to April 18, 2020. Each matrix was then normalized so that every entry *m*_*i*j_(*t*) (*i* ≠ *J*) indicates the probability an individual commuting from county *J* to county *i* on a given day. The resulting series of normalized matrices describes how inter-county connectivity changed over time during the early COVID-19 outbreak.

### The branching process model

To represent individual variation in disease transmissibility, we used a negative binomial distribution to generate the number of secondary infections (or offspring) caused by an infectious person. Denote *P*_*n*_(*X* = *x*) as the probability of an infectious person *n* generating *x* secondary infections. We define

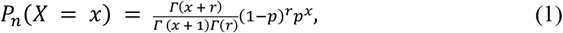

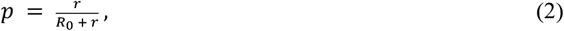

where *R*_0_ is the basic reproduction number of the disease (i.e., the average number of infections caused by one infected individual in a fully susceptible population) and *r* ∈ (0, ∞ ) is the dispersion parameter, characterizing the transmission heterogeneity among individuals. For a given basic reproduction number *R*_0_, a smaller *r* leads to a distribution *P*_*n*_ with a long tail, indicating a higher probability of producing superspreading events. Conversely, a larger *r* yields a distribution with a more homogeneous transmission. For *r* → ∞, the negative binomial distribution converges to a Poisson distribution. The distributions *P*_*n*_ with varying *r* values are shown in Fig. S5.

Model simulations proceeded as follows. For each exposed individual who contracted the disease on day *t*, we tracked the progress of disease states. Specifically, the individual became infectious on day *t* + *z*, where *z* is the latency period drawn from a Gamma distribution *Γ*_*z*_(*Z*_*a*_, *Z*_*b*_), and then recovered on day *t* + *z* + *d*, where *d* is the infectious period drawn from a Gamma distribution *Γ*_*D*_(*D*_*a*_, *D*_*b*_). The infected individual generated *k* secondary infections during the entire course of infection, computed as a random number drawn from *NB*(*R*_0_, *r*) multiplied by the fraction of susceptible population (considering the depletion of susceptible population). For each secondary infection, we assigned their infection time as day *t* + *z* + *vd*, where *v* follows a uniform distribution *U*[0,1]. That is, the timing of secondary infections was uniformly distributed within the infectious period of the primary infection. We further distributed these secondary infections spatially into different counties using commuting data. For instance, secondary infections generated in county *i* were distributed to county *J* with a probability *m*_j*i*_.

In our simulations, the latency period distribution was set as *Γ*_Z_(*Z*_*a*_ = 3, *Z*_*b*_ = 1) and the infectious period distribution was set as *Γ*_*D*_(*D*_*a*_ = 5, *D*_*b*_ = 1). For each combination of *R*_0_ and *r*, 300 independent simulations were performed. For simulations with a fixed reproduction number across space and time, we initialized the outbreak with 100 infected individuals in New York County, New York. For scenarios with spatiotemporally varying reproduction numbers, we distributed the initial infections across multiple locations to mimic geographically dispersed seeding events: 50 individuals in New York County, New York, 50 in Santa Clara County, California, 20 in Suffolk County, Massachusetts, and 20 in King County, Washington.

### Configuration of the BLL-GNN

We implemented a Bayesian Last-Layer Graph Neural Network (BLL-GNN) to infer the dispersion parameter *r* from simulated epidemic data. Each input sample contained a window of daily new infections across all counties, together with the mobility matrix. The encoder consisted of three graph convolutional layers (feature dimensions: input [number of time steps]→128→64→32), each followed by Exponential Linear Unit (ELU) activations and dropout (*p* = 0.5). Node embeddings were aggregated via global mean pooling, normalized using LayerNorm, and projected through a fully connected bottleneck (8 dimensions). The final output was computed through a Bayesian linear layer (implemented using the Blitz Bayesian framework^61^) to capture parameter uncertainty. The prior was defined as a two-component Gaussian mixture (wide component *σ*_1_ = 3.0, narrow component *σ*_2_ = 0.1, mixture weight *π* = 0.8).

The model was trained to predict log(r) using simulated outbreaks covering 21 dispersion parameter values ranging from 0.025 to 20. For each *r*, 300 realizations of outbreak data were generated. Optimization used the Adam optimizer and a fixed KL divergence weight (*β* = 3 × 10^−3^) during variational inference. All networks were implemented in PyTorch^62^ and PyTorch Geometric^63^. Further mathematical details and inference equations are provided in the Supplementary Information.

### Time-varying Bayesian graph neural network

In the real-world SARS-CoV-2 data, mobility across locations changed over time. To incorporate temporal heterogeneity in mobility changes, we extended the Bayesian GNN framework to handle time-varying mobility graphs. Each input sample consisted of daily new infections at all locations (node features, dimension N×T) and a sequence of adjacency matrices, *m*_*i*j_(*t*), describing mobility between counties across time (*t* =1,…,*T*). We implemented a temporal graph convolutional encoder, where at each time step node features were linearly transformed, propagated through normalized mobility matrices, and activated using the Exponential Linear Unit (ELU) function. Three layers of this operation, with output dimensions of 64, 64, and 32, were stacked, followed by a single LayerNorm and dropout layer. Temporal aggregation was also achieved by mean pooling across time. Then, similarly to the non-time-varying synthetic model, node embeddings were averaged to produce a graph-level summary vector and that vector was passed through a fully connected bottleneck and finally a Bayesian linear layer with the default Gaussian mixture prior (*σ*_1_ = 1.0, *σ*_2_ = 0.002, *π* = 0.5), yielding a predictive posterior over log(r).

### Generating synthetic outbreaks with county-specific *R*_0_ and underreporting

We generated synthetic outbreaks with county-specific, time-varying basic reproduction numbers. The initial reproduction number *R*_0,*ℓ*_ ∈ [2.5, 4], determined by the population size pop_*ℓ*_ of county *l*. Over time, the reproduction number *R*_*t,ℓ*_ decreases toward a floor value *R*_*floor*_ following a logistic function:

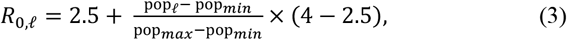

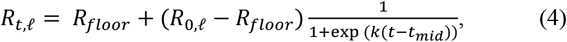

where pop_*min*_ and pop_*max*_ denote the minimum and maximum population size of all counties, *R*_*floor*_ = 1.5 is the lower bound, *k* = 0.2 is the steepness of the decay, and *t*_*mid*_ = *T*/2 is the midpoint of the simulation period *T*.

To evaluate the robustness of our inference framework under imperfect surveillance, we incorporated county-specific reporting rates *ρ*_*t,ℓ*_ for each county *l* and day *t. ρ*_*t,ℓ*_ increased monotonically over time, mimicking improving case ascertainment during the early phase of the pandemic - from 10–15% at the start to 15–20% by the end of the 60-day window. For each county *l*, the reporting rates were defined as

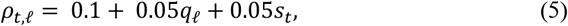

where *q*_*ℓ*_ ∼ *𝒰* (0, 1) represents the relative initial case reporting in county *ℓ* and *s*_*t*_ is a normalized sigmoid curve representing the nonlinear increase of reporting rate:

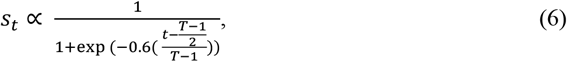

where *s*_0_ = 0 and *s*_*T*_ = 1.

### Validation of the inference framework

To validate the inference framework, we simulated one realization of an outbreak using the branching process model with a given ground-truth dispersion parameters *r* and the time-varying *R*_*t,ℓ*_, producing the daily new infections 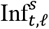 across all counties during *T* days. To estimate the crude reproduction number, we aggregated the daily county-level infections to the national level and used EpiEstim to reconstruct the national time-varying reproduction number 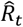 (Fig.S4). The inferred 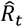 were used to generate a set of synthetic training datasets spanning a broad range of *r* values. Here we used 21 *r* values selected from 0.025 to 20. Then, we trained the BLL-GNN on these synthetic datasets and applied it to that the single realization 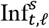 to estimate *r*. In total, we performed inference for seven ground-truth dispersion parameters. Among them, three values were not used in the training datasets, serving as out-of-sample validation targets.

When incorporating underreporting rates, we multiplied the daily infections by *ρ*_*t,ℓ*_ to obtain the corresponding observed cases *O*_*t,ℓ*_. The effective reproduction number 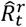 was inferred using the national observed cases using the EpiEstim. The model was then trained on the training dataset generated using the estimated 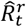. To account for the underreported infection numbers in the inputs to the BLL-GNN, we rescaled the observed cases using *O*_*t,ℓ*_/0.15, assuming a uniform, fixed reporting rate of 15% across all locations and time. This rescaling keeps the magnitude of input infections similar to that in the training datasets.

## Data Availability

All data produced are available online at https://github.com/Qing1011/branchingmodel

## Data availability

Extracted mobility network data and population data used in this paper are available at our project’s GitHub repository (https://github.com/Qing1011/branchingmodel).

## Code availability

Custom code and data supporting the statistical analysis are publicly available at GitHub (https://github.com/Qing1011/branchingmodel).

## Acknowledgements

This study was supported by funding from National Institutes of Health R35GM156799 (S.P.) and National Science Foundation DMS-2229605 (S.P.). We thank Chester Tan for discussion on graph neural network methods.

## Author contributions

S.P., Q.Y., T.B., and R.Z. conceived the study, Q.Y. performed the analysis, S.P. and Q.Y. curated the data, Q.Y., R. Z., T.B. and S.P. investigated the results. Q.Y. and S.P. drafted the manuscript, all authors revised and reviewed the manuscript.

## Supporting information

### Computation of stochastic extinction probability

The extinction probability *q*, is the probability that, starting from one infected person, the chain of transmission eventually dies out. It should be the smallest value satisfies *q* = *G*(*q*), where *G*(*s*) is the probability generating function (PGF) of a process and *s* ∈ [0,1] is the dummy variable of the PGF. The shape of *G*(*s*) depends on the basic reproduction number *R*_0_. When *R*_0_ ≤ 1, the only solution is *q* = 1, meaning the extinction is certain. When *R*_0_ > 1, a smaller solution 0 < *q* < 1 exists, representing the probability that fewer individuals are infected and eventually no new infections.

In the case of Negative Binomial distribution, the corresponding PGF is:

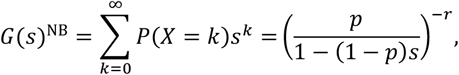

When parameterized in *R*_0_ and *r*,

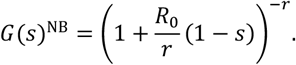

In the case of mixed Poisson distributions, the corresponding PGF is:

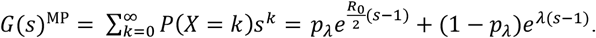

*s* = 1 is always a solution of the equation. To find other solutions for *q* = *G*(*q*)^NB^ and *q* = *G*(*q*)^MP^, we used bisection method to find the intersection of the two functions.

### Last Layer Bayesian GCN architecture and inference

We implemented a Bayesian Graph Convolutional Network (GCN) to infer the dispersion parameter *r* from simulated epidemic data. The encoder consists of three graph convolutional layers that transform node features using the normalized mobility matrix:

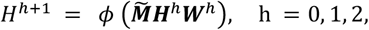

where *ϕ* is the ELU nonlinearity and 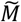 is the normalized mobility matrix. After the final convolution, node embeddings are aggregated by global mean pooling:

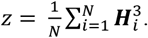

The LayerNorm and dropout (*p* = 0.5) was applied to obtain the bottleneck representation 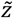. The output layer is Bayesian linear regression layer, defined as:

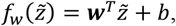

where ***w*** is the vector of weights in the last Bayesian layer, with prior

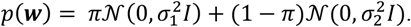

The model predicted log dispersion rate *y* and assume a Gaussian likelihood for training labels.

During the training, variational inference is performed by maximizing the evidence lower bound (ELBO):

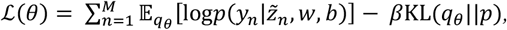

where *q*_*θ*_ is the approximate posterior distribution over the weight ***w*** and bias *b*. KL weight *β* = 3 × 10^’4^. To ensure numerical stability and to represent irreducible noise, a fixed variance floor (*σ*_min_ = 0.5) was added to the Gaussian likelihood during training. This prevents the model from becoming overconfident.

For posterior prediction, we draw *S* Monte Carlo samples from *q*_*θ*_ and compute

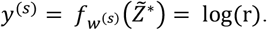

Then we get the posterior mean 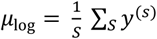 and epistemic variance *v*_ep_ = Var(*y*^(*s*)^).

To ensure numerical stability, we added a minimum value a floor sigma *σ*_min_ = 0.5 to predicted variances:

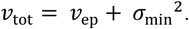

Then we assume log*r* ∼ *𝒩*(*μ*_log_, *v*_tot_), and report mean 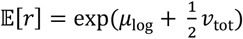, median 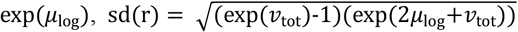 and 95% confidence interval CI_0.HI_(*r*) = vexp(*μ*_log_ − *zσ*_tot_), exp(*μ*_log_ + *zσ*_tot_)w, with *z* = 1.9599639845 and 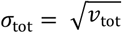.

---

**Fig. S1.**
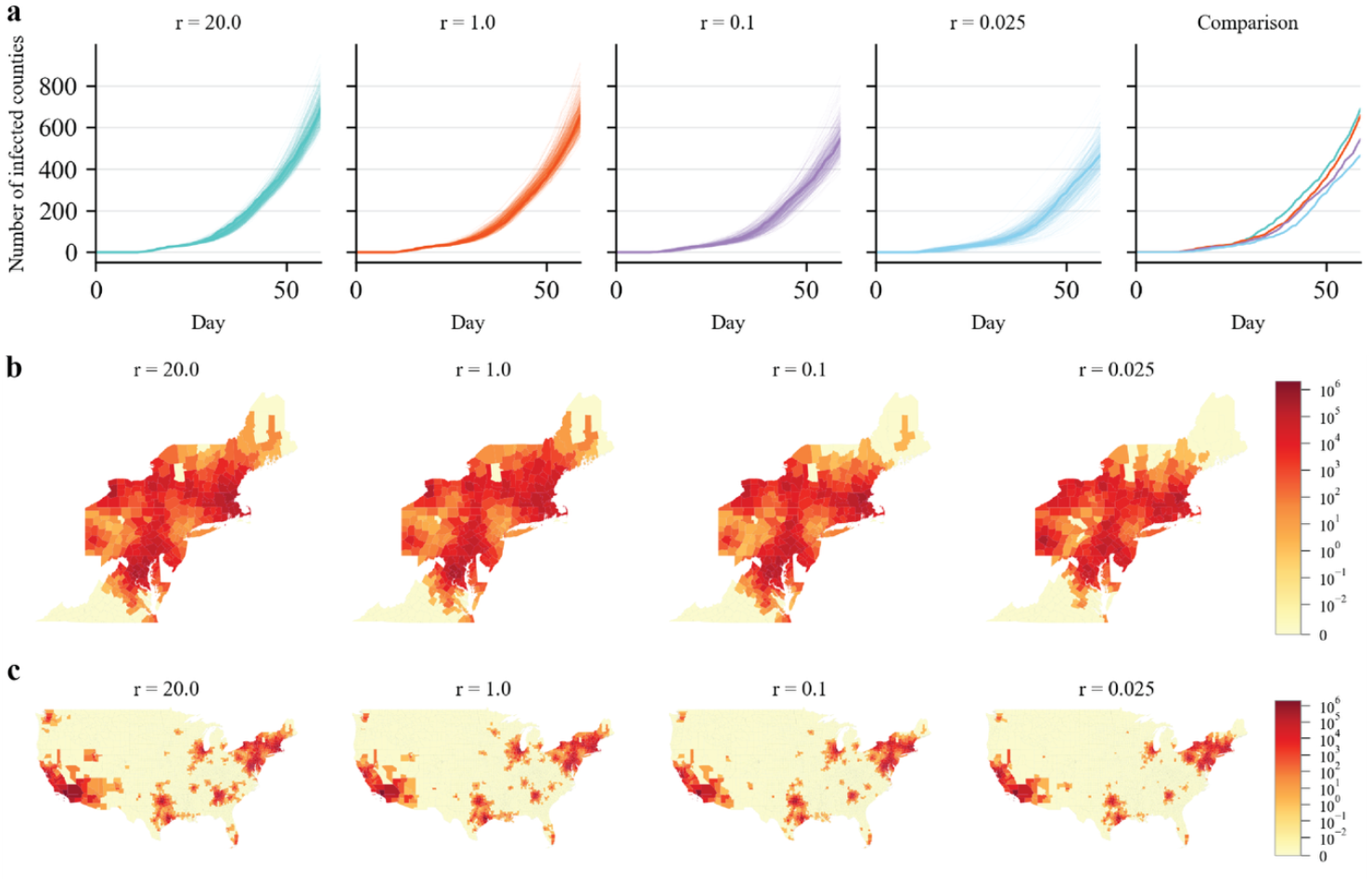
Effect of superspreading on spatial spread for *R*_0_ = 5. 5. **(a)** shows the number of counties with at least 10 daily new infections per 100,000 people over 60 days. For each parameter setting, we performed 300 independent simulations. Thick solid lines show the trajectory for the realization that infected the median number of counties on T=60 days. All the other realizations are plotted using thinner trajectories. **(b)** and **(c)** show daily new infections in each county on day 60 in the northeastern and mainland US, respectively. We visualize the simulation whose number of infected counties on day 60 is the median of the ensemble.

**Fig. S2.**
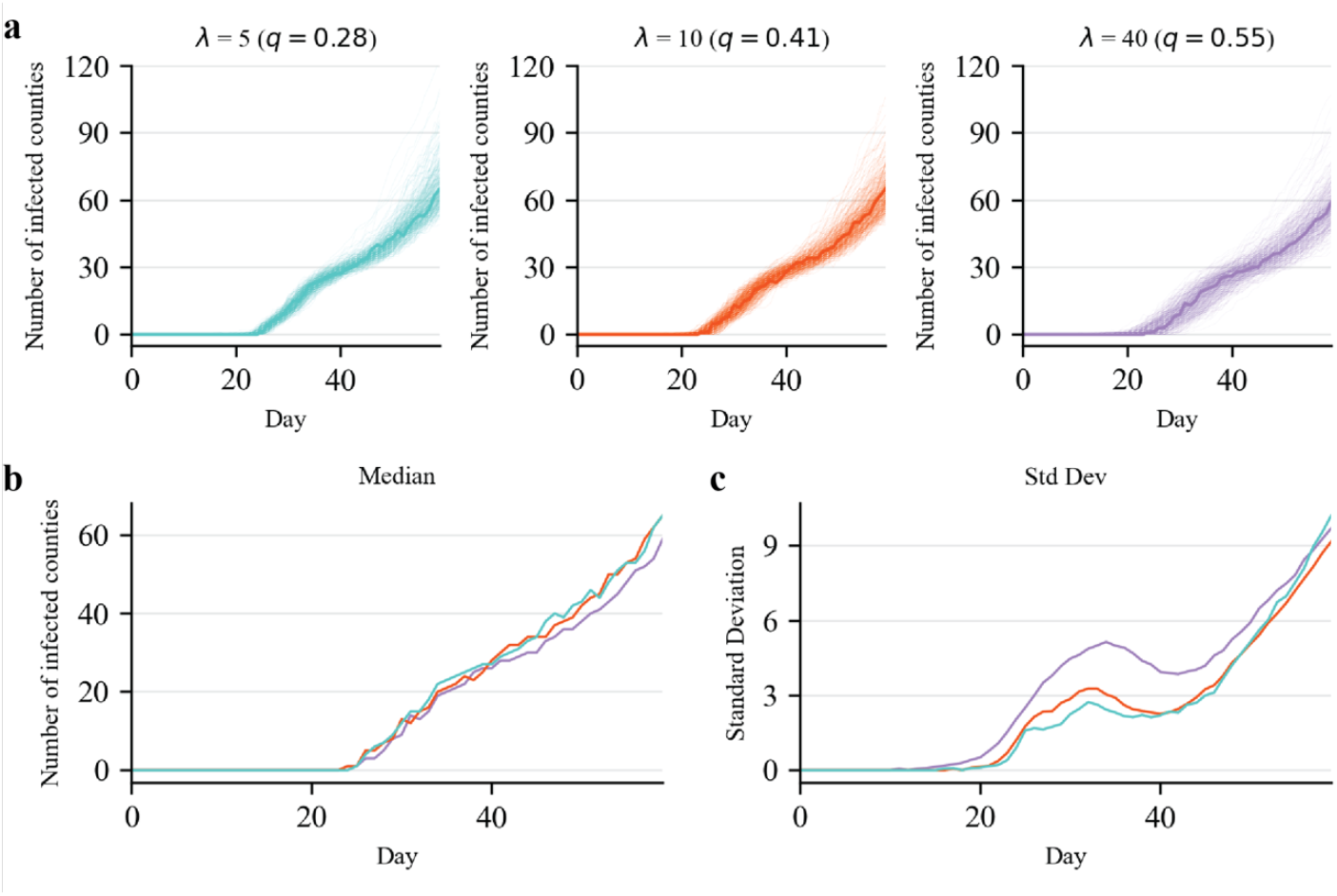
Simulations using Poisson mixture distributions for the number of secondary infections. We simulate outbreaks with *R*_0_ = 2.5 originating from New York County, starting from 100 initial infections. Simulations were performed for four different *λ* values. (**a**) The plots show the number of counties with at least 10 daily new infections per 100,000 people over 60 days. For each parameter setting, we performed 300 independent simulations. The *q* values in the parentheses are extinction probabilities for different *λ* values. Thick solid lines show the median trajectory among the 300 realizations (minimizes total L1 distance to others). All the other realizations are plotted using thinner trajectories. (**b**) Comparison of the median trajectories. (**c**) The standard deviation of number of infected counties on each day across 300 simulations.

**Fig. S3.**
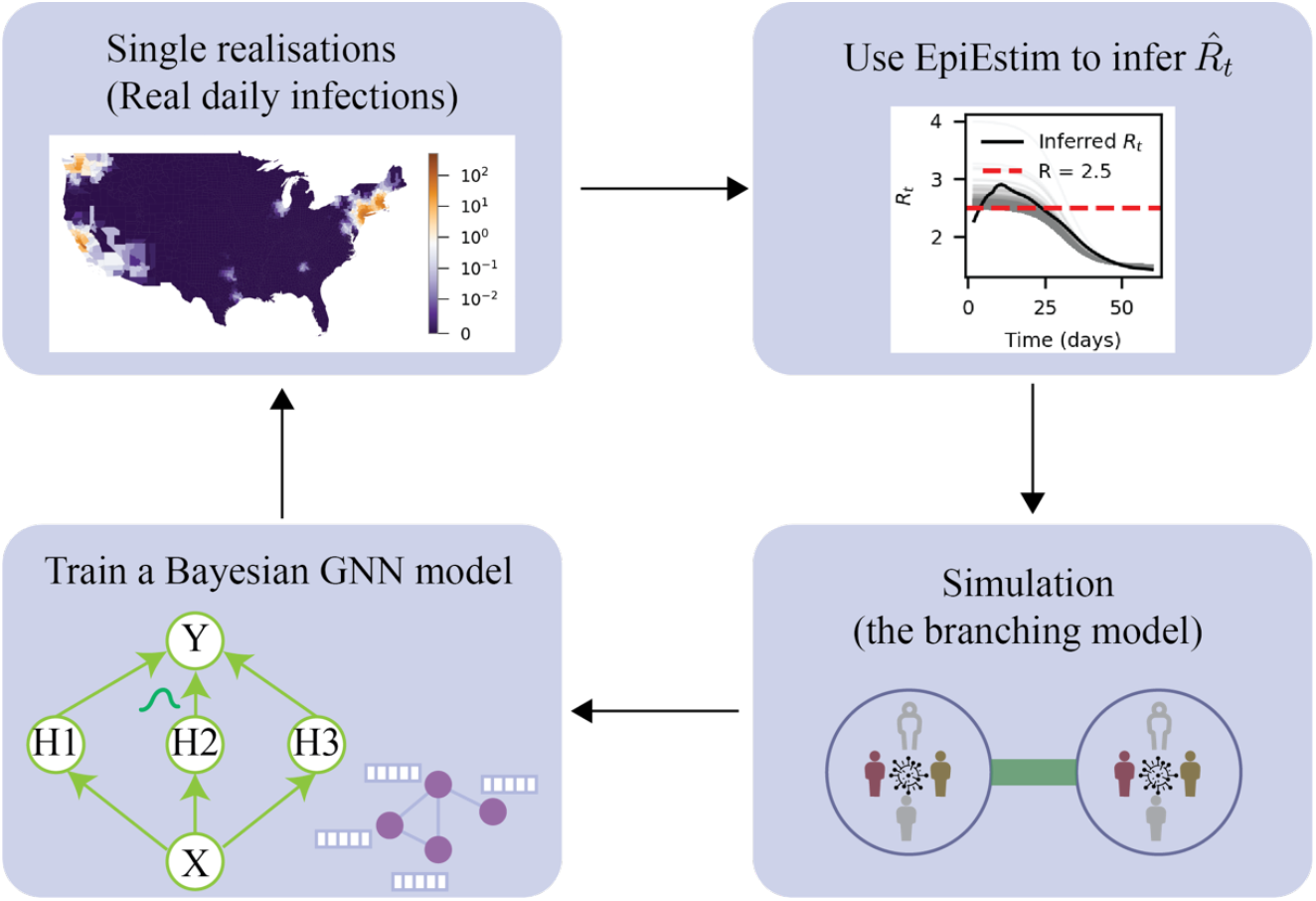
The inference pipeline. Schematic overview of the workflow for inferring the dispersion parameter *r*. Realized daily infections across US counties are first used with EpiEstim to estimate the time-varying reproduction number 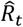. These estimates parameterize the branching-process simulation with various level of superspreading potentials. The simulated spatiotemporal infection data are then used to train a Bayesian GNN model, which learns to predict the dispersion parameter from simulated incidence data. The trained model is finally applied to the real daily infection data to infer superspreading potential.

**Fig. S4.**
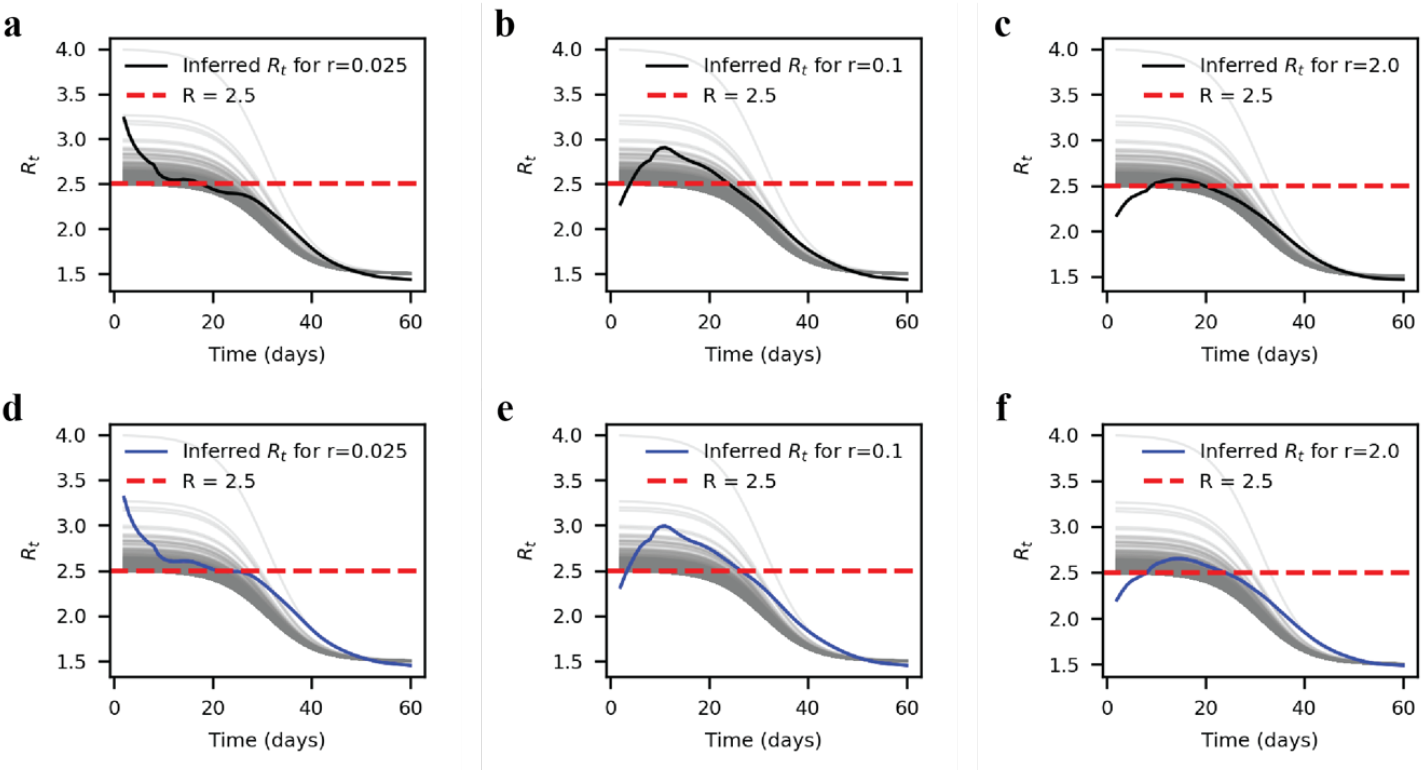
Estimated national reproduction numbers compared with location-specific reproduction numbers. Panels **(a–c)** show the national reproduction numbers estimated using EpiEstim for simulated outbreaks with dispersion parameters *r* = 0.025, *r* = 0.1 and *r* = 2.0, respectively. Grey lines indicate the actual county-specific reproduction numbers *R*_*t,ℓ*_ used in generating the outbreaks. Solid black lines represent the estimated national-level reproduction number from the aggregated daily new infections across all counties. (**d-f**). We additionally incorporated county-specific, time-varying reporting rate to generate observed daily infection data with underreporting. Solid blue lines show the reproduction number inferred from underreported observed cases. The estimated national reproduction numbers obtained from EpiEstim captured the general temporal trend of *R*_*t,ℓ*_, regardless of underreporting.

**Fig. S5.**
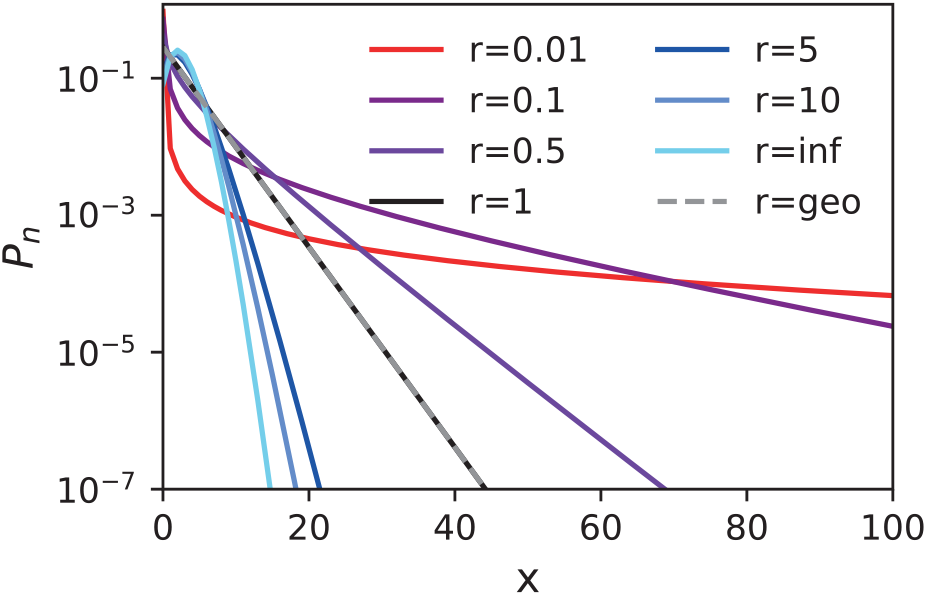
The probability density function of negative binomial distributions. The distribution of a smaller r has a longer tail and indicates a higher probability of superspreading events.

**Table S1.** Estimated basic reproduction numbers and dispersion parameters for a list of common infectious diseases.

| **Pathogens** | **Estimated basic reproduction $R_0$** | **Estimated dispersion rate $r$** | **References** |
| --- | --- | --- | --- |
| SARS-CoV-2 | 2.03–2.77 | 0.06–2.97 | [1], [2] |
| SARS | 0.94–1.88 | 0.11–0.64 | [3] |
| MERS-CoV | 0.24–1.3 | 0.045–0.14 | [4], [5] |
| Smallpox | 0.8–3.91 | 0.29–2.32 | [3] |
| Mpox | 0.21–0.42 | 0.14–3.57 | [3], [6] |
| Measles | 11–18 | 0.12–0.65 | [3], [7] |

**Table S2.** Extinction probabilities for negative binomial distributions and Poisson mixture distributions.

| Distribution | Parameter | $P(X = 0)$ | $q$ |
| --- | --- | --- | --- |
| Negative Binomial | $r = 0.025$ | 0.89 | 0.96 |
| | $r = 0.1$ | 0.72 | 0.86 |
| | $r = 1.0$ | 0.29 | 0.4 |
| | $r = 20.0$ | 0.095 | 0.13 |
| Poisson mixture | $\lambda = 200$ | 0.28 | 0.61 |
| | $\lambda = 40$ | 0.28 | 0.55 |
| | $\lambda = 10$ | 0.25 | 0.41 |
| | $\lambda = 5$ | 0.19 | 0.28 |

